# Plasma markers of neurologic injury and systemic inflammation in individuals with self-reported neurologic post-acute sequelae of SARS-CoV-2 infection (PASC)

**DOI:** 10.1101/2021.11.02.21265778

**Authors:** Michael J. Peluso, Hannah M. Sans, Carrie A. Forman, Alyssa N. Nylander, Hsi-en Ho, Scott Lu, Sarah A. Goldberg, Rebecca Hoh, Viva Tai, Sadie E. Munter, Ahmed Chenna, Brandon C. Yee, John W. Winslow, Christos J. Petropoulos, Jeffrey N. Martin, J. Daniel Kelly, Matthew S. Durstenfeld, Priscilla Y. Hsue, Peter W. Hunt, Meredith Greene, Felicia C. Chow, Joanna Hellmuth, Timothy J. Henrich, David V. Glidden, Steven G. Deeks

## Abstract

**Background:** The biologic mechanisms underlying neurologic post-acute-sequelae of SARS-CoV-2 infection (PASC) are incompletely understood.

**Methods:** We measured markers of neuronal injury (glial fibrillary acidic protein [GFAP], neurofilament light chain [NfL]) and soluble markers of inflammation among a cohort of people with prior confirmed SARS-CoV-2 infection at early and late recovery following the initial illness (defined as less than and greater than 90 days, respectively). The primary clinical outcome was the presence of self-reported central nervous system (CNS) PASC symptoms during the late recovery timepoint. We compared fold-changes in marker values between those with and without CNS PASC symptoms using linear mixed effects models and examined relationships between neurologic and immunologic markers using rank linear correlations.

**Results:** Of 121 individuals, 52 reported CNS PASC symptoms. During early recovery, those who went on to report CNS PASC symptoms had elevations in GFAP (1.3-fold higher mean ratio, 95% CI 1.04-1.63, p=0.02), but not NfL (1.06-fold higher mean ratio, 95% CI 0.89-1.26, p=0.54). During late recovery, neither GFAP nor NfL levels were elevated among those with CNS PASC symptoms. Although absolute levels of NfL did not differ, those who reported CNS PASC symptoms demonstrated a stronger downward trend over time in comparison to those who did not report CNS PASC symptoms (p=0.041). Those who went on to report CNS PASC also exhibited elevations in IL-6 (48% higher during early recovery and 38% higher during late recovery), MCP-1 (19% higher during early recovery), and TNF-alpha (19% higher during early recovery and 13% higher during late recovery). GFAP and NfL correlated with levels of several immune activation markers during early recovery; these correlations were attenuated during late recovery.

**Conclusions:** Self-reported neurologic symptoms present >90 days following SARS-CoV-2 infection are associated with elevations in markers of neurologic injury and inflammation at early recovery timepoints, suggesting that early injury can result in long-term disease. The correlation of GFAP and NfL with markers of systemic immune activation suggests one possible mechanism that might contribute to these symptoms. Additional work is needed to better characterize these processes and to identify interventions to prevent or treat this condition.

**Key Points:** *Question:* Do individuals with and without self-reported neurologic symptoms following SARS-CoV-2 infection have different levels of biomarkers of neurologic injury or immune activationã

*Findings:* In this cohort study of 121 adults, individuals reporting neurologic symptoms beyond 90 days following SARS-CoV-2 infection had higher levels of glial fibrillary acidic protein but not neurofilament light chain. Levels of several markers of inflammation including interleukin-6, tumor necrosis factor-alpha, and monocyte chemoattractant protein-1 were also elevated.

*Meaning:* Post-acute neurologic symptoms following SARS-CoV-2 infection are associated with significant differences in levels of certain biomarkers. Further investigation may provide clues to the biologic pathways underlying these symptoms.

## BACKGROUND

There is an urgent need to understand the pathophysiology that underlies the post-acute sequelae of SARS-CoV-2 infection (PASC), a condition characterized by persistent symptoms in some individuals recovering from coronavirus disease 2019 (COVID-19).^1^ While a spectrum of symptoms is reported among individuals experiencing PASC, neurologic symptoms are particularly common.^1–5^ Limited data are available on the biologic predictors and correlates of these symptoms.

Neurologic involvement during COVID-19 is common.^6–8^ Acute illness is associated with substantial immune activation^9–11^ and central nervous system (CNS) dysfunction.^11–16^ A handful of studies have measured serum glial fibrillary acidic protein (GFAP), an intermediate filament protein found in the cytoskeleton of CNS astrocytes,^17^ and neurofilament light chain (NfL), a cytoskeletal protein expressed in the axons of neurons^18^ during acute COVID-19.^19–28^ In some cases, higher levels of these markers were identified in patients with neurologic symptoms during acute infection;^21–25^ other studies have not identified such relationships.^27,28^ These markers could be of prognostic value in acute COVID-19.^23,24,29–31^

A high proportion of individuals experience ongoing physical or mental health symptoms in the 2-12 months following COVID-19.^32–37^ Recent work has suggested that immune activation might play a role in PASC.^13,38^ Inflammatory markers such as interleukin (IL)-6, tumor necrosis factor (TNF)-alpha, and interferon-gamma induced protein (IP)-10 have been associated with ongoing symptoms.^38^ These markers, particularly IL-6, may correlate with long-term neuropsychiatric manifestations of COVID-19.^14^ However, there are limited data on the relationship between biomarkers of inflammation, neurologic injury, and neurologic symptoms during the post-acute period.^25,39,40^

In a prior analysis utilizing a broad case definition of PASC (i.e., presence of any 1 of 32 COVID-19-attributed symptoms), we found that differences in levels of inflammatory markers predicted the presence of symptoms >90 days following COVID-19.^38^ In the current report, we investigated a more specific outcome defined by 5 self-reported neurologic symptoms. We evaluated markers of CNS injury and systemic inflammation among those with and without this more specific phenotype. A better understanding of the relationships between these markers among individuals with neurologic manifestations of PASC could help in identifying therapies to prevent and/or manage this condition as the pandemic continues.

## METHODS

### Participants and procedures

Beginning in April 2020 we conducted a prospective longitudinal study of individuals who had recently recovered from confirmed SARS-CoV-2 infection (Long-term Impact of Infection with Novel Coronavirus (LIINC) cohort; NCT04362150).^41^ The vast majority (78%) had not been hospitalized during the acute phase. The determination of eligibility was agnostic to the presence or absence of persistent SARS-CoV-2 attributed symptoms.

A research coordinator administered a study questionnaire at early (=<90 days) and late (>90 days) recovery time points following COVID-19 symptom onset. Participants were queried regarding the presence of 32 symptoms, including 8 neurologic symptoms (Supplemental Table 1). A symptom was recorded as present if it was reported at the time of the visit and was either new in onset since the time of SARS-CoV-2 infection or had worsened since the time of SARS-CoV-2 infection. Symptoms that preceded and were unchanged following SARS-CoV-2 infection were recorded as absent. We also collected information about demographics and medical history, and retrospectively collected information on symptoms experienced during the acute phase of the illness. Blood was collected by venipuncture at each visit.

### Clinical outcomes

The primary clinical outcome was central nervous system (CNS) PASC, defined as the presence of at least one CNS symptom at a late recovery visit occurring >90 days from initial COVID-19 symptom onset. These symptoms included: memory/concentration issues, headache, vision problems, dizziness, and balance issues. We selected these symptoms because they were felt to best reflect dysfunction of the central nervous system and most likely to associate with biologic processes that could be identified using the two primary biomarker outcomes. A secondary analysis examined any neurological symptom, which included the following in addition to the central neurological symptoms: problems with smell or taste, smelling an odor that is not really present, and numbness/tingling (Supplemental Table 2).

### Biomarker assays

Plasma biomarker measurements were performed using the fully automated HD-X Simoa platform at two timepoints: early recovery (median 52 days) and late recovery (median 123 days). Those performing the assays were blinded to clinical information. The primary analytes were plasma GFAP and NfL measured using the GFAP Discovery and NF-light Advantage kit assays, respectively (Quanterix). We also measured levels of markers that have been found to be important during acute SARS-CoV-2 infection^10,11^ using multiplex (Cytokine 3-PlexA: IL-6, IL-10, TNF-alpha) or single-plex (IFNγ, IP-10, MCP-1) kits. SARS-CoV-2 receptor binding domain (RBD) IgG was also assayed. All assays were performed according to the manufacturer’s instructions and assay performance was consistent with the manufacturer’s specifications.

### Statistical analysis

We log-transformed all biomarkers to reduce influence of outliers and to permit interpretation of fold-changes. As in prior work,^38^ we compared the ratio of the mean transformed values for each biomarker between those with and without persistent neurologic symptoms using linear mixed effects models with terms for PASC, time period (early versus late recovery), and their interaction. This approach permits comparison of the values at early and late time points as well as assessment of whether trajectories in marker values differ between those with and without persistent symptoms. We calculated fold-changes and 95% confidence intervals by exponentiating the coefficients to give the ratio between the untransformed biomarker values. We used Spearman correlations to evaluate relationships between levels of neurologic and immune markers. All p-values are two-sided. We used Stata (version 16.1; StataCorp, College Station, TX) and Prism (version 9.1.2, GraphPad Software, L.L.C., San Diego, CA).

### Ethics

All participants provided written informed consent. The study was approved by the Institutional Review Board at the University of California, San Francisco.

## RESULTS

### Study participants

The study included 121 individuals with primary outcome data (Table 1), 92 of whom (76%) had a paired early recovery sample for analysis. During acute COVID-19, the majority (89, 74%) had been symptomatic outpatients, while 27 (22%) had been hospitalized. Five (4%) reported asymptomatic SARS-CoV-2 infection. Of those hospitalized, 23 (85%) required supplemental oxygen and 3 (11%) required mechanical ventilation. SARS-CoV-2-targeted treatment was uncommon: 4 individuals (15%) received remdesivir, 1 (4%) received convalescent plasma, and 5 (19%) received steroids. No hospitalized participant was known to have experienced an acute neurological event during their hospitalization. All samples were collected prior to the availability of SARS-CoV-2 vaccination.

**Table 1:**
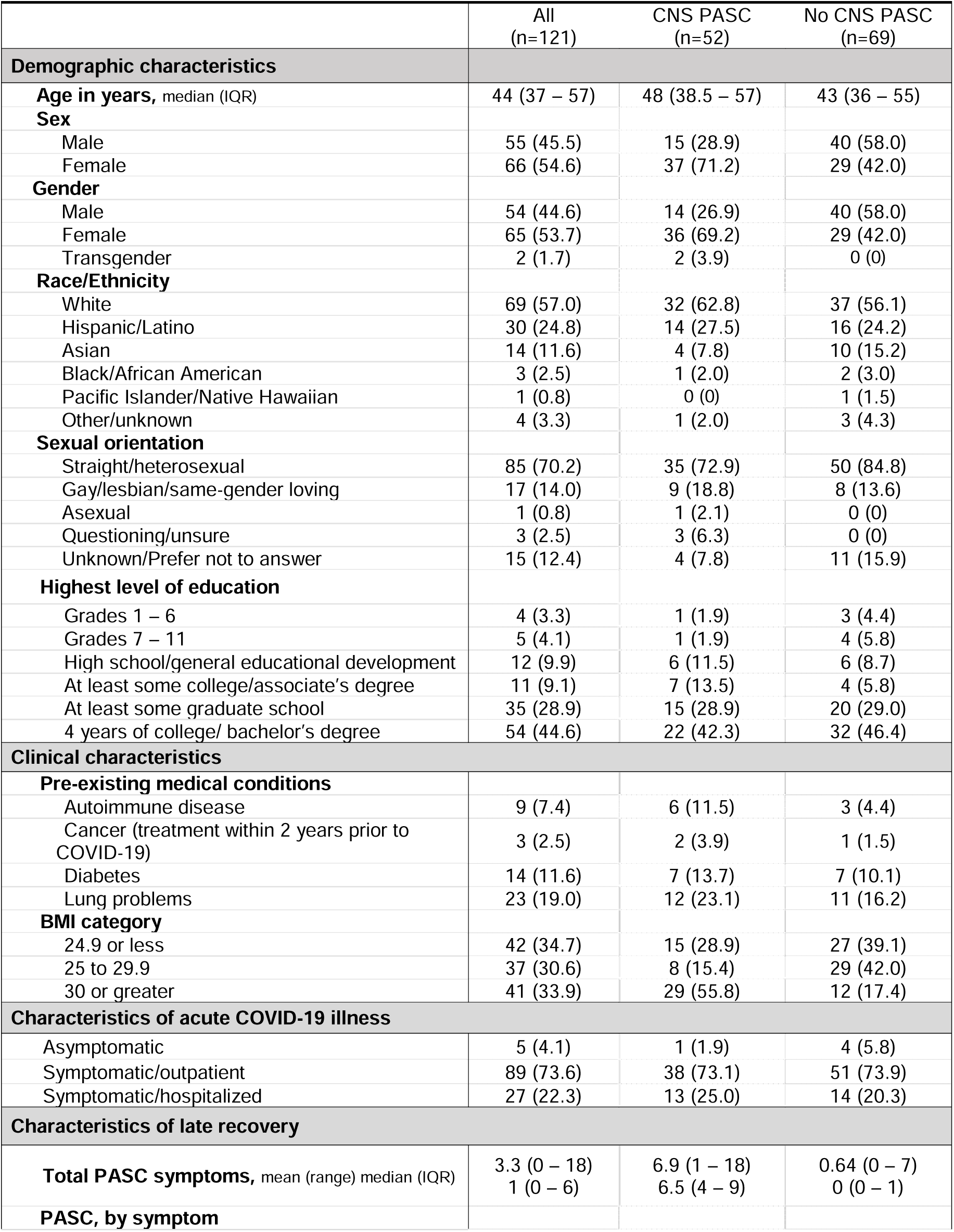
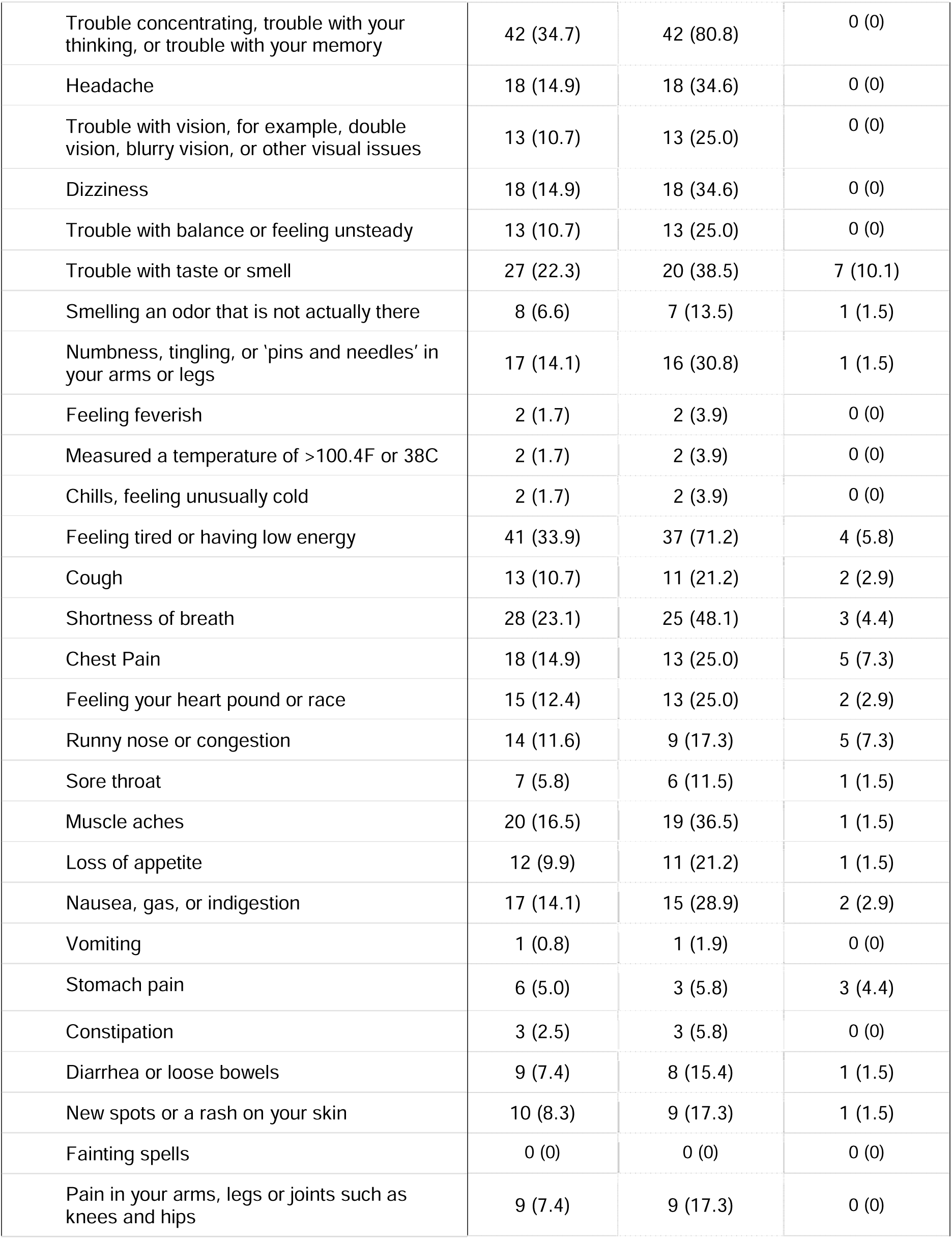
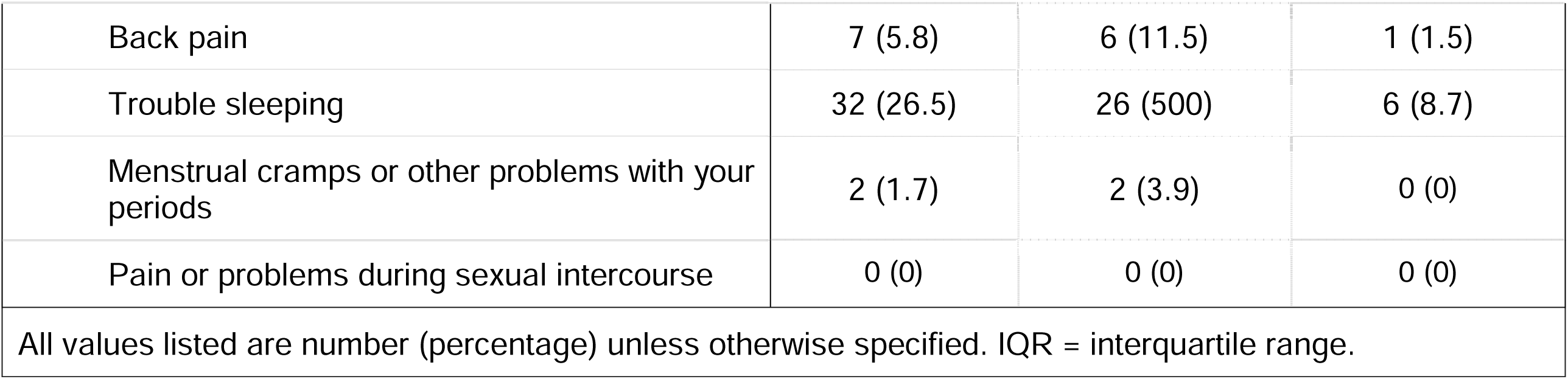
Characteristics of study cohort.

|  | All<br>(n=121) | CNS PASC<br>(n=52) | No CNS PASC<br>(n=69) |
| --- | --- | --- | --- |
| **Demographic characteristics** |  |  |  |
| **Age in years**, median (IQR) | 44 (37 – 57) | 48 (38.5 – 57) | 43 (36 – 55) |
| **Sex** |  |  |  |
| Male | 55 (45.5) | 15 (28.9) | 40 (58.0) |
| Female | 66 (54.6) | 37 (71.2) | 29 (42.0) |
| **Gender** |  |  |  |
| Male | 54 (44.6) | 14 (26.9) | 40 (58.0) |
| Female | 65 (53.7) | 36 (69.2) | 29 (42.0) |
| Transgender | 2 (1.7) | 2 (3.9) | 0 (0) |
| **Race/Ethnicity** |  |  |  |
| White | 69 (57.0) | 32 (62.8) | 37 (56.1) |
| Hispanic/Latino | 30 (24.8) | 14 (27.5) | 16 (24.2) |
| Asian | 14 (11.6) | 4 (7.8) | 10 (15.2) |
| Black/African American | 3 (2.5) | 1 (2.0) | 2 (3.0) |
| Pacific Islander/Native Hawaiian | 1 (0.8) | 0 (0) | 1 (1.5) |
| Other/unknown | 4 (3.3) | 1 (2.0) | 3 (4.3) |
| **Sexual orientation** |  |  |  |
| Straight/heterosexual | 85 (70.2) | 35 (72.9) | 50 (84.8) |
| Gay/lesbian/same-gender loving | 17 (14.0) | 9 (18.8) | 8 (13.6) |
| Asexual | 1 (0.8) | 1 (2.1) | 0 (0) |
| Questioning/unsure | 3 (2.5) | 3 (6.3) | 0 (0) |
| Unknown/Prefer not to answer | 15 (12.4) | 4 (7.8) | 11 (15.9) |
| **Highest level of education** |  |  |  |
| Grades 1 – 6 | 4 (3.3) | 1 (1.9) | 3 (4.4) |
| Grades 7 – 11 | 5 (4.1) | 1 (1.9) | 4 (5.8) |
| High school/general educational development | 12 (9.9) | 6 (11.5) | 6 (8.7) |
| At least some college/associate's degree | 11 (9.1) | 7 (13.5) | 4 (5.8) |
| At least some graduate school | 35 (28.9) | 15 (28.9) | 20 (29.0) |
| 4 years of college/ bachelor's degree | 54 (44.6) | 22 (42.3) | 32 (46.4) |
| **Clinical characteristics** |  |  |  |
| **Pre-existing medical conditions** |  |  |  |
| Autoimmune disease | 9 (7.4) | 6 (11.5) | 3 (4.4) |
| Cancer (treatment within 2 years prior to COVID-19) | 3 (2.5) | 2 (3.9) | 1 (1.5) |
| Diabetes | 14 (11.6) | 7 (13.7) | 7 (10.1) |
| Lung problems | 23 (19.0) | 12 (23.1) | 11 (16.2) |
| **BMI category** |  |  |  |
| 24.9 or less | 42 (34.7) | 15 (28.9) | 27 (39.1) |
| 25 to 29.9 | 37 (30.6) | 8 (15.4) | 29 (42.0) |
| 30 or greater | 41 (33.9) | 29 (55.8) | 12 (17.4) |
| **Characteristics of acute COVID-19 illness** |  |  |  |
| Asymptomatic | 5 (4.1) | 1 (1.9) | 4 (5.8) |
| Symptomatic/outpatient | 89 (73.6) | 38 (73.1) | 51 (73.9) |
| Symptomatic/hospitalized | 27 (22.3) | 13 (25.0) | 14 (20.3) |
| **Characteristics of late recovery** |  |  |  |
| **Total PASC symptoms**, mean (range) median (IQR) | 3.3 (0 – 18)<br>1 (0 – 6) | 6.9 (1 – 18)<br>6.5 (4 – 9) | 0.64 (0 – 7)<br>0 (0 – 1) |
| **PASC, by symptom** |  |  |  |
| Trouble concentrating, trouble with your thinking, or trouble with your memory | 42 (34.7) | 42 (80.8) | 0 (0) |
| Headache | 18 (14.9) | 18 (34.6) | 0 (0) |
| Trouble with vision, for example, double vision, blurry vision, or other visual issues | 13 (10.7) | 13 (25.0) | 0 (0) |
| Dizziness | 18 (14.9) | 18 (34.6) | 0 (0) |
| Trouble with balance or feeling unsteady | 13 (10.7) | 13 (25.0) | 0 (0) |
| Trouble with taste or smell | 27 (22.3) | 20 (38.5) | 7 (10.1) |
| Smelling an odor that is not actually there | 8 (6.6) | 7 (13.5) | 1 (1.5) |
| Numbness, tingling, or 'pins and needles' in your arms or legs | 17 (14.1) | 16 (30.8) | 1 (1.5) |
| Feeling feverish | 2 (1.7) | 2 (3.9) | 0 (0) |
| Measured a temperature of >100.4F or 38C | 2 (1.7) | 2 (3.9) | 0 (0) |
| Chills, feeling unusually cold | 2 (1.7) | 2 (3.9) | 0 (0) |
| Feeling tired or having low energy | 41 (33.9) | 37 (71.2) | 4 (5.8) |
| Cough | 13 (10.7) | 11 (21.2) | 2 (2.9) |
| Shortness of breath | 28 (23.1) | 25 (48.1) | 3 (4.4) |
| Chest Pain | 18 (14.9) | 13 (25.0) | 5 (7.3) |
| Feeling your heart pound or race | 15 (12.4) | 13 (25.0) | 2 (2.9) |
| Runny nose or congestion | 14 (11.6) | 9 (17.3) | 5 (7.3) |
| Sore throat | 7 (5.8) | 6 (11.5) | 1 (1.5) |
| Muscle aches | 20 (16.5) | 19 (36.5) | 1 (1.5) |
| Loss of appetite | 12 (9.9) | 11 (21.2) | 1 (1.5) |
| Nausea, gas, or indigestion | 17 (14.1) | 15 (28.9) | 2 (2.9) |
| Vomiting | 1 (0.8) | 1 (1.9) | 0 (0) |
| Stomach pain | 6 (5.0) | 3 (5.8) | 3 (4.4) |
| Constipation | 3 (2.5) | 3 (5.8) | 0 (0) |
| Diarrhea or loose bowels | 9 (7.4) | 8 (15.4) | 1 (1.5) |
| New spots or a rash on your skin | 10 (8.3) | 9 (17.3) | 1 (1.5) |
| Fainting spells | 0 (0) | 0 (0) | 0 (0) |
| Pain in your arms, legs or joints such as knees and hips | 9 (7.4) | 9 (17.3) | 0 (0) |
| Back pain | 7 (5.8) | 6 (11.5) | 1 (1.5) |
| Trouble sleeping | 32 (26.5) | 26 (500) | 6 (8.7) |
| Menstrual cramps or other problems with your periods | 2 (1.7) | 2 (3.9) | 0 (0) |
| Pain or problems during sexual intercourse | 0 (0) | 0 (0) | 0 (0) |
| All values listed are number (percentage) unless otherwise specified. IQR = interquartile range. |  |  |  |

Fifty-two individuals, of whom the majority were women, reported CNS symptoms at the late recovery timepoint (Table 1). Among them, the most commonly reported CNS symptoms were trouble concentrating (81%) headache (35%) and dizziness (35%).

Early recovery visits took place at a median of 52 (IQR 38-64) days post-infection. Late recovery visits took place at a median of 123 (IQR 114-135) days post-infection. The early recovery visit for those reporting CNS PASC symptoms occurred slightly later than for those who denied CNS PASC symptoms (60 [IQR 40-67] and 49 [IQR 37-59] days, respectively). The timing of the late recovery visit was similar between those with and without CNS PASC (123 [IQR 117-137] and 124 [IQR 113-134] days, respectively).

### Levels of biomarkers among those with and without CNS PASC symptoms

We first compared levels of each marker measured during early recovery between those who went on to report CNS PASC symptoms and those who did not (Figure 1, Supplemental Table 3). At the early recovery timepoint, those who went on to report CNS PASC had significantly higher levels of GFAP (1.3-fold higher mean ratio, 95% CI 1.04-1.63, p=0.02), but not NfL (1.06-fold higher mean ratio, 95% CI 0.89-1.26, p=0.54). Those who went on to report CNS PASC also had higher levels of IL-6 (1.48-fold higher mean ratio, 95% CI 1.12-1.96, p=0.006), MCP-1 (1.19-fold higher mean ratio, 95% CI 1.01-1.40, p=0.034), and TNF-alpha (1.19-fold higher mean ratio, 95% CI 1.06-1.34, p=0.003) compared to those who did not report CNS PASC. Trends for other markers were in a similar direction, although the differences did not achieve statistical significance.

**Figure 1.**
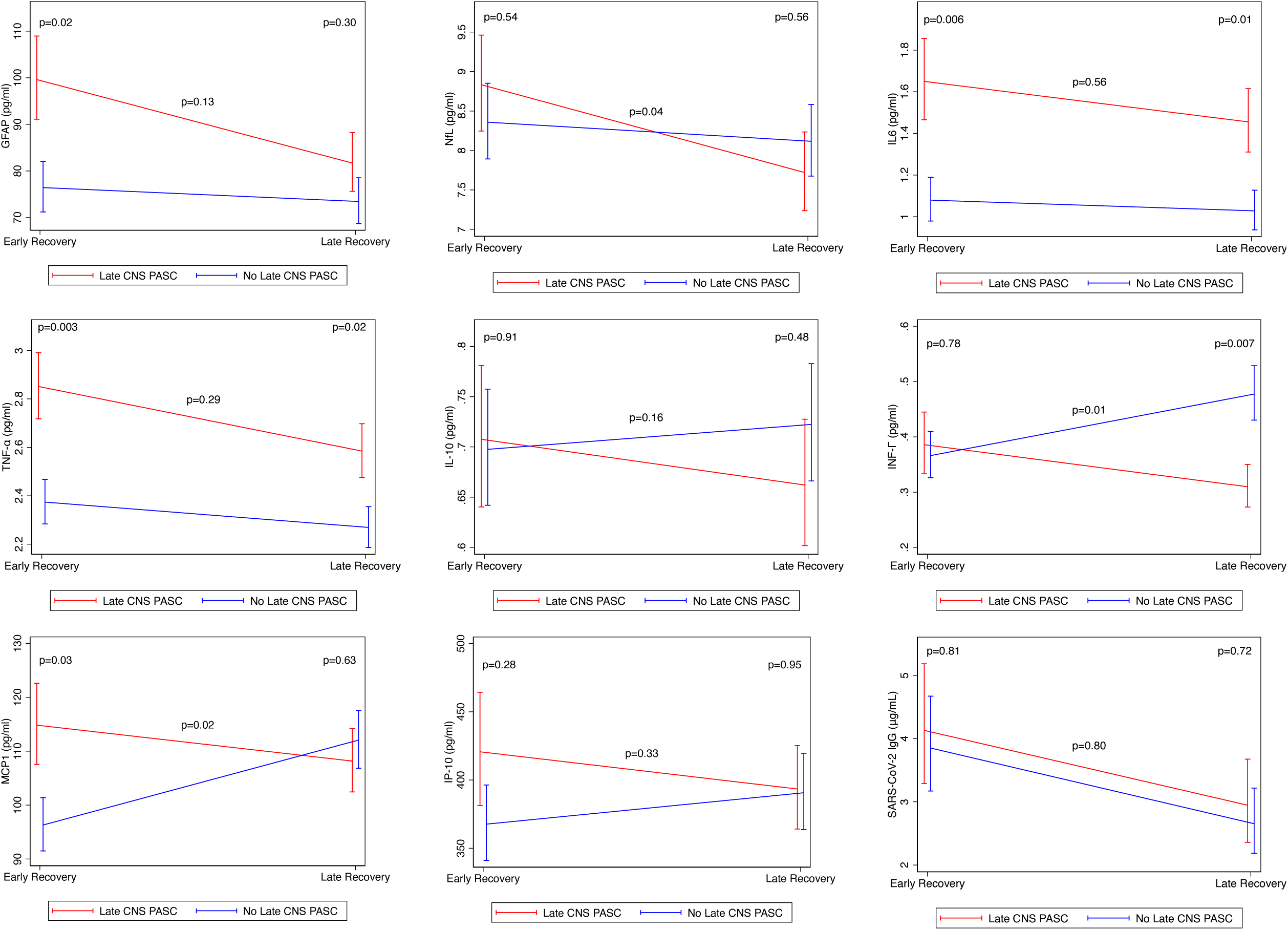
Cross-sectional measurements and longitudinal trends in biomarker levels among those with and without CNS PASC. P-values reflect group comparisons during early and late recovery, as well as comparison of change over time between groups. GFAP, glial fibrillary acidic protein; NfL, neurofilament light chain; IL-6, interleukin-6; TNF-alpha, tumor necrosis factor alpha; IL-10, interleukin-10; IFN-gamma, interferon-gamma; MCP-1, monocyte chemoattractant protein 1; IP-10, interferon-gamma induced protein 10; SARS-CoV-2, severe acute respiratory syndrome coronavirus 2; IgG immunoglobulin G. Early recovery represents a median of 52 days post-SARS-CoV-2 symptom onset (or positive PCR); late recovery represents a median of 123 days post-SARS-COV-2 symptom onset (or positive PCR).

We next compared levels of each biomarker measured during late recovery between those with and without self-reported CNS PASC at this visit (Figure 1, Supplemental Table 3). No significant differences were detected in GFAP or NfL between those with and without PASC (Figure 1). Those reporting persistent CNS PASC symptoms had persistent elevations in IL-6 (1.38-fold higher mean ratio, 95% CI 1.07-1.77, p=0.013), and TNF-alpha (1.13-fold higher mean ratio, 95% CI 1.02-1.26, p=0.022). Interferon-gamma was lower (0.71-fold difference, 95% CI 0.55-0.91, p=0.007). Levels of SARS-CoV-2 RBD IgG did not differ between groups at either the early or late timepoints.

### Changes in levels of biomarkers over time

To examine changes in the levels of these markers between the early and late recovery timepoints, we used mixed models to indicate changes over time among those with and without CNS PASC symptoms (Figure 1, Supplemental Table 3). Significant differences in trends of NFL (p=0.041), MCP-1 (p=0.019), and IFN-gamma (p=0.012) were noted between the CNS PASC and non-CNS PASC groups. As predicted from the cross-sectional analyses, consistently higher levels of IL-6 and TNF-alpha were observed, although the trends in the levels of these markers did not differ between groups.

### Relationships between neurologic and inflammatory markers

To examine relationships between the neurological markers and markers of inflammation, we performed nonparametric pairwise analyses at early and late recovery timepoints (Figure 2). GFAP levels weakly correlated with MCP-1 (r=0.21, p=0.02) and IL-6 (r=0.18, p=0.054) at the early timepoint and with IL-6 at the late timepoint (r=0.19, p=0.043). NfL correlated with MCP-1 (r=0.41, p<0.001), IL-6 (r=0.23, p=0.012), IFN-gamma (r=0.28, p=0.003), and TNF-alpha (r=0.32, p<0.001) at the early timepoint and with MCP-1 (r=0.31, p<0.001) at the late timepoint. In addition, there was a strong correlation between NfL and SARS-CoV-2 IgG at the early timepoint (r=0.40, p<0.001).

**Figure 2.**
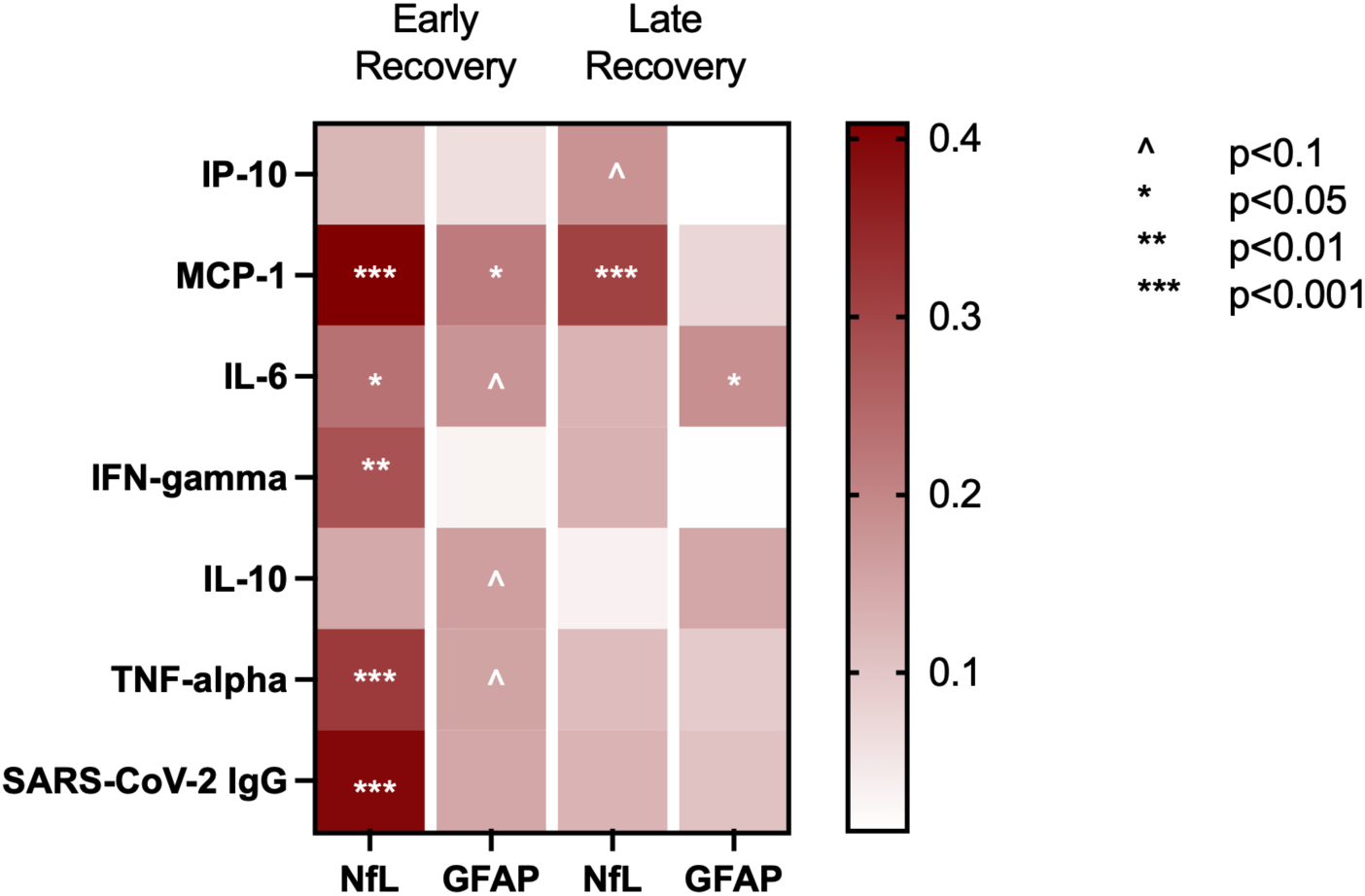
Relationships between biomarkers of neurologic injury and systemic inflammation in the full cohort. Data reflect non-parametric correlations between markers at early and late recovery timepoints. GFAP, glial fibrillary acidic protein; NfL, neurofilament light chain; IL-6, interleukin-6; TNF-alpha, tumor necrosis factor alpha; IL-10, interleukin-10; IFN-gamma, interferon-gamma; MCP-1, monocyte chemoattractant protein 1; IP-10, interferon-gamma induced protein 10; SARS-CoV-2, severe acute respiratory syndrome coronavirus 2; IgG immunoglobulin G. Early recovery represents a median of 52 days post-SARS-CoV-2 symptom onset (or positive PCR); late recovery represents a median of 123 days post-SARS-COV-2 symptom onset (or positive PCR).

### Influence of symptoms during acute infection

We did not identify significant differences in levels of markers at either recovery timepoint between those with and without prior CNS symptoms during acute infection (Supplemental Table 4). For some markers, we noted non-significant trends toward differential changes over time in groups with and without CNS symptoms during acute infection. These included NfL (more steep decline among those with acute CNS symptoms, p=0.066) and anti-RBD IgG (less steep decline among those with acute CNS symptoms, p=0.063).

### Sensitivity analyses

Because of the relationship between age and levels of NfL,^42^ we performed an age-adjusted analysis which did not change the primary results (no new relationship between NfL and PASC was identified (Supplemental Table 5). We also repeated the primary analysis adjusting for age, sex, and prior hospitalization status which did not change the interpretation of the results (Supplemental Table 6), although some of the relationships were slightly attenuated.

We performed a secondary analysis in which individuals reporting any symptom that could be attributed to a primary neurologic cause (including the peripheral nervous system) were compared against individuals reporting no neurologic symptoms (Supplemental Table 7). In this analysis, the elevation in GFAP seen at early follow-up among those reporting any neurologic PASC symptom was slightly attenuated (mean ratio 1.24, 95% CI 1.00-1.55, p=0.052). Similar elevations were seen in IL-6, MCP-1, and TNF-alpha at early follow-up among those reporting any neurologic PASC symptom. At late follow-up, those with any neurologic PASC had higher levels of IL-6, and TNF-alpha. The difference in interferon-gamma seen among those with CNS PASC in the primary analysis was no longer statistically significant.

Finally, because we have previously found that PASC is associated with elevations in certain markers, we performed an analysis comparing those with CNS PASC to those reporting no symptoms of any kind during late recovery (Supplemental Table 8). The interpretation of the primary results was again unchanged, although some of the findings were attenuated.

## DISCUSSION

A large proportion of individuals with PASC experience symptoms that may be attributed to nervous system dysfunction,^1–5^ but the pathophysiologic processes underlying such symptoms remain poorly understood. We investigated associations between self-reported neurologic symptoms and plasma biomarkers of CNS injury and systemic inflammation during early and late recovery periods following laboratory-confirmed SARS-CoV-2 infection. Those reporting CNS PASC symptoms approximately four months following initial infection had earlier elevations in several biomarkers, including GFAP, IL-6, and TNF-alpha suggesting that the acute infection resulted in direct CNS tissue injury and systemic inflammation, both of which might be causally related to the development of CNS PASC. Replication of these findings in larger and more diverse cohorts may be a first step toward identifying interventions for their prevention and/or management.

Elevations in GFAP during early recovery were associated with later CNS PASC. While we did not observe elevations in NfL at either timepoint, we did identify a more precipitous decline among those reporting CNS PASC. Together, these observations lend support to the possibility of early injury that resolves while clinical symptoms persist. Our findings are in line with studies that identified a correlation between NfL and/or GFAP with neurologic symptoms during the acute phase, but found no association between the biomarker levels and persistence of neurologic symptoms after six months.^39,40^ Further work exploring the dynamics of these markers in cohorts with measurements performed during the period of acute illness, as well as efforts to identify downstream markers which may persist during later recovery, will be informative.

This analysis builds upon several observations we and others have made suggesting that markers of systemic inflammation may be important in driving PASC.^13,38^ While we previously observed associations between such markers and broadly defined PASC (i.e., any 1 of 32 COVID-19-attributed symptoms),^38^ it is notable that the strength of these associations was more pronounced in the current analysis utilizing a more specific PASC outcome (i.e., any 1 of 5 CNS PASC symptoms). At the same time, the fact that those reporting CNS PASC symptoms had a greater number of PASC symptoms overall makes it challenging to disentangle a more specific CNS PASC phenotype from more severe PASC in general. It suggests that CNS PASC might reflect one extreme on a spectrum of illness. Further work to compare those with distinct phenotypic clusters of symptoms, if they exist, could further elucidate the biology.

Dysregulation of IL-6 and TNF-alpha is potentially deleterious in inflammatory disease states,^43,44^ related to systemic and localized tissue inflammation and endothelial dysfunction. The reason for elevations in levels of these markers among those with CNS PASC is not clear. One possibility is that they represent residual inflammation from acute infection that is slower to resolve among those with PASC. However, the identification of persistent differences months following infection suggests other possibilities such as a delayed return to immunologic homeostasis related to ongoing pathophysiology (e.g., persistent antigenic stimulation,^45^ microvascular dysfunction,^46–48^ autoimmunity,^49^ etc.). While the source of these inflammatory markers is unknown, both IL-6 and TNF-alpha can be produced by CNS as well as peripheral immune cells and both cytokines have been implicated in CNS pathology.^50,51^ Similarly, MCP-1 is a chemokine expressed by macrophages and microglia, and elevations have been implicated in neurocognitive conditions.^52,53^ Further investigation into the source of IL-6, TNF-alpha, and MCP-1 among those with CNS PASC, which may include co-investigation of the peripheral blood and CSF compartment, will be needed to provide clues to the pathophysiology underlying this condition.

Interestingly, levels of these biomarkers during early and late recovery did not differ between those with and without self-reported neurologic symptoms during acute infection. This suggests that residual neurologic injury in those with neuro-symptomatic acute COVID-19, at least among study populations comprised primarily of outpatients, is not the most important causal factor. The determination of who will have ongoing elevations in markers of neurologic injury and/or who will go on to experience CNS PASC during late recovery appears to be more complex than identifying those who report neurologic symptoms during the acute phase of illness.

While the pathogenesis of neurologic complications from SARS-CoV-2 is yet unclear, several hypotheses have been proposed. These include direct viral infection, systemic inflammation, compartmentalized neuroinflammation, and sequelae of thrombotic injury. Viral tropism for human astrocytes has been demonstrated in-vivo,^54^ post-mortem brain samples from COVID-19 patients have shown preferential infection of astrocytes,^55^ and a case-control study of brain samples uncovered altered gene expression in some astrocytes.^56^ Astrocyte dysfunction could relate to emerging cognitive PASC complaints, which can encapsulate attention and working memory deficits.^57^ Direct invasion of neurons has been suggested based on their expression of ACE-2 receptor, but SARS-Cov-2 viral particles have been only rarely demonstrated in neuropathological autopsy studies and studies of CSF during acute infection.^58–60^ Additionally, there is evidence of multifocal inflammatory infiltrates consisting of lymphocytes as well as activated innate immune cells in autopsy tissue.^61^ Further investigation of PASC will require in-depth exploration of what is occurring in the CNS compartment during both acute and recovery time periods.

Our analysis has several important limitations. First, while recruitment was agnostic to the presence of persistent symptoms, the cohort is not representative of the general population with PASC. Second, we relied on self-report to ascertain the presence of symptoms. This risks misattribution of symptoms to neurologic causes and therefore misclassification of individuals as having CNS PASC. In addition, it is difficult to disentangle neurologic symptoms from non-neurologic symptoms which might co-occur and it is possible that differences are driven by more severe PASC in general rather than neurologic symptoms specifically. Affective symptoms may also co-occur and be inter-related. Third, we did not include any objective neurologic measurements, and studies that do include such measurements (which may include detailed neurological history and examination, neurocognitive and neuropsychiatric testing, and/or neuroimaging) are likely to be more informative. Fourth, we measured a limited set of biomarkers. Our measurements were all taken in blood, and while there are established relationships between blood and CSF measurements in other disease conditions,^62,63^ these have yet to be established for COVID-19.^28^ For this reason, more detailed studies that include CSF analyses will be critical. Finally, paired pre-pandemic specimens were not available, and it is possible that elevations in these markers among those with CNS PASC preceded SARS-CoV-2 infection. Regardless, we believe that the observations made here provide important clues to important biological pathways to inform more detailed neurological evaluations and potential therapeutic studies.

## Data Availability

All data produced in the present work are available upon reasonable request to the authors

## FOOTNOTES

## Acknowledgements

We are grateful to the LIINC study participants and to the clinical staff who provided care to these individuals during their acute illness period and during their recovery. We thank Dr. Isabel Rodriguez-Barraquer, Dr. Bryan Greenhouse, and Dr. Rachel Rutishauser for their contributions to the LIINC leadership team. We acknowledge current and former LIINC clinical study team members Tamara Abualhsan, Andrea Alvarez, Mireya Arreguin, Melissa Buitrago, Monika Deswal, Emily Fehrman, Heather Hartig, Yanel Hernandez, Marian Kerbleski, James Lombardo, Enrique Martinez, Lynn Ngo, Dylan Ryder, Ruth Diaz Sanchez, Cassandra Thanh, Fatima Ticas, Leonel Torres, and Meghann Williams; and LIINC laboratory team members Amanda Buck, Tyler-Marie Deveau, Joanna Donatelli, Jill Hakim, Nikita Iyer, Owen Janson, Christopher Nixon, Isaac Thomas, and Keirstinne Turcios. We thank Elnaz Eilkhani for coordination with the Institutional Review Board. We acknowledge the contributions of the UCSF Clinical and Translational Science Unit, Core Immunology Laboratory, and AIDS Specimen Bank. We thank Jeremy Lambert from Quanterix for his advice and assistance with the SARS-CoV-2 IgG antibody assay kits.

## Author Contributions

MJP, HH, JNM, JDK, MG, FCC, JH, TJH, and SGD designed the study, which was supported through funding to MJP, JDK, TJH, and SGD. MJP, ANN, RH, VT, and JDK collected clinical data and biospecimens, which were processed by SEM in the laboratory of TJH. Specimens were analyzed by AC, BCY, JWW, and CJP. MJP, SL, SAG, and DVG performed and/or interpreted the statistical analyses. MJP, HMS, CAF, ANN, HH, JNM, DVG, and SGD drafted the initial manuscript with input from JDK, AC, BCY, JWW, CJP, PWH, MSD, PYH, MG, TJH, FCC, and JH. All authors edited, reviewed, and approved the final manuscript.

