## Supplemental Tables for "Plasma markers of neurologic injury and systemic inflammation in individuals with self-reported neurologic post-acute sequelae of SARS-CoV-2 infection (PASC)"

**Supplemental Table 1. Queried symptoms and associated categorization.**

| **Symptom query** | **Category** |
| --- | --- |
| Trouble concentrating, trouble with your thinking, or trouble with your memory | Central neurologic symptom |
| Headache | Central neurologic symptom |
| Trouble with vision, for example, double vision, blurry vision, or other visual issues | Central neurologic symptom |
| Dizziness | Central neurologic symptom |
| Trouble with balance or feeling unsteady | Central neurologic symptom |
| Trouble with taste or smell | Other neurologic symptom |
| Smelling an odor that is not actually there | Other neurologic symptom |
| Numbness, tingling, or ‘pins and needles’ in your arms or legs | Other neurologic symptom |
| Feeling feverish | Non-neurologic symptom |
| Measured a temperature of >100.4F or 38C | Non-neurologic symptom |
| Chills, feeling unusually cold | Non-neurologic symptom |
| Cough | Non-neurologic symptom |
| Shortness of breath | Non-neurologic symptom |
| Chest Pain | Non-neurologic symptom |
| Feeling your heart pound or race | Non-neurologic symptom |
| Runny nose or congestion | Non-neurologic symptom |
| Sore throat | Non-neurologic symptom |
| Muscle aches | Non-neurologic symptom |
| Loss of appetite | Non-neurologic symptom |
| Nausea, gas, or indigestion | Non-neurologic symptom |
| Vomiting | Non-neurologic symptom |
| Stomach pain | Non-neurologic symptom |
| Constipation | Non-neurologic symptom |
| Diarrhea or loose bowels | Non-neurologic symptom |
| New spots or a rash on your skin | Non-neurologic symptom |
| Fainting spells | Non-neurologic symptom |
| Back pain | Non-neurologic symptom |
| Menstrual cramps or other problems with your periods | Non-neurologic symptom |
| Pain or problems during sexual intercourse | Non-neurologic symptom |

| **Supplemental Table 2: Additional characteristics of study cohort.** | | | | | | |
| --- | --- | --- | --- | --- | --- | --- |
|  | All  (n=121) | CNS PASC (n=52) | No CNS PASC (n=69) | Any Neuro PASC (n=59) | Any PASC (n=73) | No PASC (n=48) |
| **Demographic characteristics** |  |  |  |  |  |  |
| **Age in years,** median (IQR) | 44 (37 – 57) | 48 (38.5 – 57) | 43 (36 – 55) | 48 (34 – 57) | 44 (36 – 56) | 44.5 (37 – 58.5) |
| **Sex** |  |  |  |  |  |  |
| Male | 55 (45.5) | 15 (28.9) | 40 (58.0) | 19 (32.2) | 28 (38.4) | 27 (56.3) |
| Female | 66 (54.6) | 37 (71.2) | 29 (42.0) | 40 (67.8) | 45 (61.6) | 21 (43.8) |
| **Gender** |  |  |  |  |  |  |
| Male | 54 (44.6) | 14 (26.9) | 40 (58.0) | 18 (30.5) | 27 (37.0) | 27 (56.3) |
| Female | 65 (53.7) | 36 (69.2) | 29 (42.0) | 39 (66.1) | 44 (60.3) | 21 (43.8) |
| Transgender | 2 (1.7) | 2 (3.9) | 0 (0) | 2 (3.4) | 2 (2.7) | 0 (0) |
| **Race/Ethnicity** |  |  |  |  |  |  |
| White | 69 (57.0) | 32 (62.8) | 37 (56.1) | 36 (62.1) | 41 (57.8) | 28 (60.9) |
| Hispanic/Latino | 30 (24.8) | 14 (27.5) | 16 (24.2) | 16 (27.6) | 22 (31.0) | 8 (17.4) |
| Asian | 14 (11.6) | 4 (7.8) | 10 (15.2) | 5 (8.6) | 6 (8.5) | 8 (17.4) |
| Black/African American | 3 (2.5) | 1 (2.0) | 2 (3.0) | 1 (1.7) | 2 (2.8) | 1 (2.2) |
| Pacific Islander/Native Hawaiian | 1 (0.8) | 0 (0) | 1 (1.5) | 0 (0) | 0 (0) | 1 (2.2) |
| Other/unknown | 4 (3.3) | 1 (2.0) | 3 (4.3) | 1 (1.7) | 2 (2.8) | 2 (4.2) |
| **Sexual orientation** |  |  |  |  |  |  |
| Straight/heterosexual | 85 (70.2) | 35 (72.9) | 50 (84.8) | 42 (76.4) | 52 (76.5) | 33 (84.6) |
| Gay/lesbian/same-gender loving | 17 (14.0) | 9 (18.8) | 8 (13.6) | 9 (16.4) | 12 (17.7) | 5 (12.8) |
| Asexual | 1 (0.8) | 1 (2.1) | 0 (0) | 1 (1.8) | 1 (1.5) | 0 (0) |
| Questioning/unsure | 3 (2.5) | 3 (6.3) | 0 (0) | 3 (5.5.) | 3 (4.4) | 0 (0) |
| Unknown/Prefer not to answer | 15 (12.4) | 4 (7.8) | 11 (15.9) | 4 (6.8) | 5 (6.8) | 10 (20.8) |
| **Highest level of education** |  |  |  |  |  |  |
| Grades 1 – 6 | 4 (3.3) | 1 (1.9) | 3 (4.4) | 1 (1.7) | 1 (1.4) | 3 (6.3) |
| Grades 7 – 11 | 5 (4.1) | 1 (1.9) | 4 (5.8) | 2 (3.4) | 3 (4.1) | 2 (4.2) |
| High school/general educational development | 12 (9.9) | 6 (11.5) | 6 (8.7) | 6 (10.2) | 8 (11.0) | 4 (8.3) |
| At least some college/associate’s degree | 11 (9.1) | 7 (13.5) | 4 (5.8) | 7 (11.9) | 10 (13.7) | 1 (2.1) |
| At least some graduate school | 35 (28.9) | 15 (28.9) | 20 (29.0) | 17 (28.8) | 21 (28.8) | 14 (29.2) |
| 4 years of college/ bachelor’s degree | 54 (44.6) | 22 (42.3) | 32 (46.4) | 26 (44.1) | 30 (41.1) | 24 (50.0) |
| **Clinical characteristics** | | | | | | |
| **Pre-existing medical conditions** |  |  |  |  |  |  |
| Autoimmune disease | 9 (7.4) | 6 (11.5) | 3 (4.4) | 6 (10.2) | 8 (11.0) | 1 (2.1) |
| Cancer (treatment within 2 years prior to COVID-19) | 3 (2.5) | 2 (3.9) | 1 (1.5) | 2 (3.5) | 2 (2.8) | 1 (2.1) |
| Diabetes | 14 (11.6) | 7 (13.7) | 7 (10.1) | 7 (12.1) | 8 (11.1) | 6 (12.5) |
| Lung problems | 23 (19.0) | 12 (23.1) | 11 (16.2) | 13 (22.0) | 13 (18.1) | 10 (20.8) |
| **BMI category** |  |  |  |  |  |  |
| 24.9 or less | 42 (34.7) | 15 (28.9) | 27 (39.1) | 18 (30.5) | 25 (34.3) | 17 (35.4) |
| 25 to 29.9 | 37 (30.6) | 8 (15.4) | 29 (42.0) | 12 (20.3) | 18 (24.7) | 19 (39.6) |
| 30 or greater | 41 (33.9) | 29 (55.8) | 12 (17.4) | 29 (49.2) | 30 (41.1) | 11 (22.9) |
| **Characteristics of acute COVID-19 illness** | | | | | | |
| Asymptomatic | 5 (4.1) | 1 (1.9) | 4 (5.8) | 1 (1.7) | 2 (2.7) | 3 (6.3) |
| Symptomatic/outpatient | 89 (73.6) | 38 (73.1) | 51 (73.9) | 44 (74.6) | 52 (71.2) | 37 (77.1) |
| **Symptomatic/hospitalized** | 27 (22.3) | 13 (25.0) | 14 (20.3) | 14 (23.7) | 19 (26.0) | 8 (16.7) |
| Required O2 | 23 (85.2) | 12 (92.3) | 11 (78.6) | 13 (92.9) | 17 (89.5) | 6 (75.0) |
| Required ventilator | 3 (11.1) | 1 (7.7) | 2 (14.3) | 2 (14.3) | 2 (10.5) | 1 (12.5) |
| Treated with remdesivir | 4 (14.8) | 3 (23.1) | 1 (7.1) | 4 (28.6) | 4 (21.1) | 0 (0) |
| Treated with convalescent plasma | 1 (3.7) | 0 (0) | 1 (7.1) | 1 (7.1) | 1 (5.3) | 0 (0) |
| Treated with steroids | 5 (18.5) | 5 (38.4) | 0 (0) | 5 (35.7) | 5 (26.3) | 0 (0) |
| **Characteristics of Late Recovery** | | | | | | |
| **Total PASC symptoms,** mean (range) median (IQR) | 3.3 (0 – 18)  1 (0 – 6) | 6.9 (1 – 18)  6.5 (4 – 9) | 0.64 (0 – 7)  0 (0 – 1) | 6.4 (1 – 18)  6 (4 – 9) | 5.5 (1 – 18)  5 (2 – 8) | - |
| **PASC, by symptom** |  |  |  |  |  |  |
| Trouble concentrating, trouble with your thinking, or trouble with your memory | 42 (34.7) | 42 (80.8) | 0 (0) | 42 (71.2) | 42 (57.5) | 0 (0) |
| Headache | 18 (14.9) | 18 (34.6) | 0 (0) | 18 (30.5) | 18 (24.7) | 0 (0) |
| Trouble with vision, for example, double vision, blurry vision, or other visual issues | 13 (10.7) | 13 (25.0) | 0 (0) | 13 (22.0) | 13 (17.8) | 0 (0) |
| Dizziness | 18 (14.9) | 18 (34.6) | 0 (0) | 18 (30.5) | 18 (24.7) | 0 (0) |
| Trouble with balance or feeling unsteady | 13 (10.7) | 13 (25.0) | 0 (0) | 13 (22.0) | 13 (17.8) | 0 (0) |
| Trouble with taste or smell | 27 (22.3) | 20 (38.5) | 7 (10.1) | 27 (45.8) | 27 (37.0) | 0 (0) |
| Smelling an odor that is not actually there | 8 (6.6) | 7 (13.5) | 1 (1.5) | 8 (13.6) | 8 (11.0) | 0 (0) |
| Numbness, tingling, or ‘pins and needles’ in your arms or legs | 17 (14.1) | 16 (30.8) | 1 (1.5) | 17 (28.8) | 17 (23.3) | 0 (0) |
| Feeling feverish | 2 (1.7) | 2 (3.9) | 0 (0) | 2 (3.4) | 2 (2.7) | 0 (0) |
| Measured a temperature of >100.4F or 38C | 2 (1.7) | 2 (3.9) | 0 (0) | 2 (3.4) | 2 (2.7) | 0 (0) |
| Chills, feeling unusually cold | 2 (1.7) | 2 (3.9) | 0 (0) | 2 (3.4) | 2 (2.7) | 0 (0) |
| Feeling tired or having low energy | 41 (33.9) | 37 (71.2) | 4 (5.8) | 39 (66.1) | 41 (56.2) | 0 (0) |
| Cough | 13 (10.7) | 11 (21.2) | 2 (2.9) | 12 (20.3) | 13 (17.8) | 0 (0) |
| Shortness of breath | 28 (23.1) | 25 (48.1) | 3 (4.4) | 25 (42.4) | 28 (38.4) | 0 (0) |
| Chest Pain | 18 (14.9) | 13 (25.0) | 5 (7.3) | 15 (25.4) | 18 (24.7) | 0 (0) |
| Feeling your heart pound or race | 15 (12.4) | 13 (25.0) | 2 (2.9) | 14 (23.7) | 15 (20.6) | 0 (0) |
| Runny nose or congestion | 14 (11.6) | 9 (17.3) | 5 (7.3) | 10 (17.0) | 14 (19.2) | 0 (0) |
| Sore throat | 7 (5.8) | 6 (11.5) | 1 (1.5) | 6 (10.2) | 7 (9.6) | 0 (0) |
| Muscle aches | 20 (16.5) | 19 (36.5) | 1 (1.5) | 20 (33.9) | 20 (27.4) | 0 (0) |
| Loss of appetite | 12 (9.9) | 11 (21.2) | 1 (1.5) | 11 (18.6) | 12 (16.4) | 0 (0) |
| Nausea, gas, or indigestion | 17 (14.1) | 15 (28.9) | 2 (2.9) | 15 (25.4) | 17 (23.3) | 0 (0) |
| Vomiting | 1 (0.8) | 1 (1.9) | 0 (0) | 1 (1.7) | 1 (1.4) | 0 (0) |
| Stomach pain | 6 (5.0) | 3 (5.8) | 3 (4.4) | 4 (6.8) | 6 (8.2) | 0 (0) |
| Constipation | 3 (2.5) | 3 (5.8) | 0 (0) | 3 (5.1) | 3 (4.1) | 0 (0) |
| Diarrhea or loose bowels | 9 (7.4) | 8 (15.4) | 1 (1.5) | 8 (13.6) | 9 (12.3) | 0 (0) |
| New spots or a rash on your skin | 10 (8.3) | 9 (17.3) | 1 (1.5) | 9 (15.3) | 10 (13.7) | 0 (0) |
| Fainting spells | 0 (0) | 0 (0) | 0 (0) | 0 (0) | 0 (0) | 0 (0) |
| Pain in your arms, legs or joints such as knees and hips | 9 (7.4) | 9 (17.3) | 0 (0) | 9 (15.3) | 9 (12.3) | 0 (0) |
| Back pain | 7 (5.8) | 6 (11.5) | 1 (1.5) | 6 (10.2) | 7 (9.6) | 0 (0) |
| Trouble sleeping | 32 (26.5) | 26 (500) | 6 (8.7) | 26 (44.1) | 32 (43.8) | 0 (0) |
| Menstrual cramps or other problems with your periods | 2 (1.7) | 2 (3.9) | 0 (0) | 2 (3.4) | 2 (2.7) | 0 (0) |
| Pain or problems during sexual intercourse | 0 (0) | 0 (0) | 0 (0) | 0 (0) | 0 (0) | 0 (0) |
| All values listed are number (percentage) unless otherwise specified. IQR = interquartile range. | | | | | | |

**Supplemental Table 3. Primary analysis: any CNS symptom vs. no CNS symptom at late recovery.**

| Any CNS Symptom v. No CNS Symptom | | | | | |
| --- | --- | --- | --- | --- | --- |
|  | P-value | | Fold Change in Mean 95% CI | | P-value |
|  | Early Recovery | Late Recovery | Early Recovery | Late Recovery | Change over Time |
| IL-6 | 0.006 | 0.013 | 1.48 (1.12-1.96) | 1.38 (1.07-1.77) | 0.561 |
| GFAP | 0.02 | 0.298 | 1.30 (1.04-1.63) | 1.11 (0.91-1.36) | 0.134 |
| NFL | 0.537 | 0.558 | 1.06 (0.89-1.26) | 0.95 (0.81-1.12) | 0.041 |
| MCP-1 | 0.034 | 0.625 | 1.19 (1.01-1.40) | 0.97 (0.84-1.11) | 0.019 |
| IFN-gamma | 0.776 | 0.007 | 1.04 (0.78-1.39) | 0.71 (0.55-0.91) | 0.012 |
| IL-10 | 0.913 | 0.483 | 1.01 (0.81-1.26) | 0.93 (0.75-1.15) | 0.163 |
| TNF-alpha | 0.003 | 0.022 | 1.19 (1.06-1.34) | 1.13 (1.02-1.26) | 0.291 |
| IP-10 | 0.277 | 0.946 | 1.14 (0.90-1.46) | 1.01 (0.82-1.24) | 0.325 |
| SARS-CoV-2 IgG | 0.814 | 0.722 | 1.07 (0.60-1.90) | 1.11 (0.63-1.93) | 0.8 |

**Supplemental Table 4. Any CNS symptom vs. no CNS symptom during acute infection.**

|  | P-value | | Fold Change in Mean 95% CI | | P-value |
| --- | --- | --- | --- | --- | --- |
|  | Early Recovery | Late Recovery | Early Recovery | Late Recovery | Change over Time |
| IL-6 | 0.94 | 0.518 | 0.99 (0.72-1.35) | 1.10 (0.82-1.48) | 0.422 |
| GFAP | 0.969 | 0.521 | 1.00 (0.77-1.28) | 1.08 (0.86-1.36) | 0.484 |
| NFL | 0.649 | 0.567 | 1.05 (0.86-1.27) | 0.95 (0.78-1.14) | 0.066 |
| MCP-1 | 0.869 | 0.908 | 0.99 (0.82-1.18) | 0.99 (0.84-1.17) | 0.957 |
| IFN-gamma | 0.368 | 0.556 | 1.16 (0.84-1.60) | 1.09 (0.81-1.47) | 0.747 |
| IL-10 | 0.482 | 0.528 | 1.09 (0.85-1.40) | 1.08 (0.85-1.38) | 0.87 |
| TNF-alpha | 0.834 | 0.877 | 1.01 (0.89-1.16) | 1.01 (0.89-1.15) | 0.937 |
| IP-10 | 0.798 | 0.499 | 1.04 (0.79-1.35) | 0.92 (0.73-1.17) | 0.411 |
| SARS-CoV-2 IgG | 0.909 | 0.371 | 1.04 (0.54-1.99) | 1.34 (0.71-2.53) | 0.063 |

**Supplemental Table 5. Any CNS symptom v. no CNS symptom at late recovery, adjusted for age.**

| Any Central Neuro Symptom v. No CNS Symptom | | | | | |
| --- | --- | --- | --- | --- | --- |
| Adjusted for Age | | | | | |
|  | P-value | | Fold Change in Mean 95% CI | | P-value |
|  | Early Recovery | Late Recovery | Early Recovery | Late Recovery | Change over Time |
| IL-6 | 0.009 | 0.014 | 1.43 (1.09-1.88) | 1.36 (1.06-1.73) | 0.675 |
| GFAP | 0.037 | 0.371 | 1.26 (1.01-1.55) | 1.09 (0.90-1.32) | 0.18 |
| NFL | 0.859 | 0.223 | 1.01 (0.88-1.16) | 0.92 (0.81-1.05) | 0.068 |
| MCP-1 | 0.063 | 0.535 | 1.16 (0.99-1.37) | 0.96 (0.83-1.10) | 0.029 |
| IFN-gamma | 0.845 | 0.006 | 1.03 (0.77-1.37) | 0.71 (0.55-0.91) | 0.015 |
| IL-10 | 0.901 | 0.49 | 1.01 (0.81-1.27) | 0.93 (0.75-1.15) | 0.161 |
| TNF-alpha | 0.004 | 0.024 | 1.18 (1.05-1.33) | 1.13 (1.02-1.25) | 0.332 |
| IP-10 | 0.363 | 0.951 | 1.12 (0.88-1.43) | 0.99 (0.81-1.22) | 0.353 |
| SARS-CoV-2 IgG | 0.88 | 0.776 | 1.04 (0.59-1.84) | 1.08 (0.62-1.88) | 0.775 |

**Supplemental Table 6. Any CNS symptom v. no CNS symptom at late recovery, adjusted for age, sex, and hospitalization status.**

| Any Central Neuro Symptom v. None | | | | | |
| --- | --- | --- | --- | --- | --- |
| Adjusted for Age, Sex and Hospitalizaton | | | | | |
|  | P-value | | Fold Change in Mean 95% CI | | P-value |
|  | Early Recovery | Late Recovery | Early Recovery | Late Recovery | Change over Time |
| IL-6 | 0.018 | 0.033 | 1.38 (1.06-1.80) | 1.30 (1.02-1.65) | 0.624 |
| GFAP | 0.083 | 0.626 | 1.21 (0.98-1.51) | 1.05 (0.86-1.28) | 0.171 |
| NFL | 0.938 | 0.141 | 0.99 (0.86-1.15) | 0.90 (0.79-1.03) | 0.063 |
| MCP-1 | 0.039 | 0.677 | 1.18 (1.01-1.39) | 0.97 (0.84-1.12) | 0.027 |
| IFN-gamma | 0.914 | 0.007 | 1.02 (0.76-1.36) | 0.70 (0.54-0.91) | 0.015 |
| IL-10 | 0.949 | 0.39 | 0.99 (0.79-1.25) | 0.91 (0.73-1.13) | 0.157 |
| TNF-alpha | <0.001 | 0.003 | 1.22 (1.09-1.37) | 1.17 (1.05-1.29) | 0.328 |
| IP-10 | 0.356 | 0.884 | 1.12 (0.88-1.44) | 0.98 (0.80-1.22) | 0.307 |
| SARS-CoV-2 IgG | 0.843 | 0.926 | 0.95 (0.56-1.60) | 0.98 (0.59-1.62) | 0.821 |

**Supplemental Table 7. Any neuro symptom v. no neuro symptom at late recovery.**

| Any Neuro Symptom v. No Neuro Symptom. | | | | | |
| --- | --- | --- | --- | --- | --- |
|  | P-value | | Fold Change in Mean 95% CI | | P-value |
|  | Early Recovery | Late Recovery | Early Recovery | Late Recovery | Change over Time |
| IL-6 | 0.005 | 0.04 | 1.48 (1.12-1.94) | 1.30 (1.01-1.67) | 0.304 |
| GFAP | 0.052 | 0.486 | 1.24 (1.00-1.55) | 1.07 (0.88-1.31) | 0.153 |
| NFL | 0.222 | 0.741 | 1.11 (0.94-1.32) | 1.03 (0.87-1.21) | 0.115 |
| MCP-1 | 0.039 | 0.584 | 1.18 (1.01-1.39) | 0.96 (0.84-1.11) | 0.019 |
| IFN-gamma | 0.391 | 0.214 | 1.13 (0.85-1.50) | 0.85 (0.66-1.10) | 0.063 |
| IL-10 | 0.104 | 0.526 | 1.20 (0.96-1.49) | 1.07 (0.87-1.32) | 0.068 |
| TNF-alpha | 0.002 | 0.03 | 1.19 (1.06-1.34) | 1.12 (1.01-1.25) | 0.211 |
| IP-10 | 0.191 | 0.849 | 1.17 (0.92-1.48) | 0.98 (0.80-1.20) | 0.159 |
| SARS-CoV-2 IgG | 0.833 | 0.484 | 1.06 (0.60-1.87) | 1.22 (0.70-2.11) | 0.271 |

**Supplemental Table 8. Any CNS symptom v. no PASC symptom at late recovery.**

| Any CNS Symptom v. No PASC Symptom | | | | | |
| --- | --- | --- | --- | --- | --- |
|  | P-value | | Fold Change in Mean 95% CI | | P-value |
|  | Early Recovery | Late Recovery | Early Recovery | Late Recovery | Change over Time |
| IL-6 | 0.007 | 0.002 | 1.47 (1.11-1.95) | 1.50 (1.16-1.94) | 0.9 |
| GFAP | 0.059 | 0.674 | 1.28 (0.99-1.65) | 1.05 (0.84-1.32) | 0.113 |
| NFL | 0.519 | 0.716 | 1.06 (0.89-1.27) | 0.97 (0.81-1.15) | 0.085 |
| MCP-1 | 0.056 | 0.582 | 1.20 (1.00-1.44) | 0.96 (0.81-1.12) | 0.029 |
| IFN-gamma | 0.391 | 0.227 | 1.13 (0.86-1.48) | 0.87 (0.69-1.09) | 0.095 |
| IL-10 | 0.245 | 0.868 | 1.10 (0.94-1.30) | 1.01 (0.87-1.18) | 0.201 |
| TNF-alpha | 0.003 | 0.048 | 1.20 (1.06-1.36) | 1.12 (1.00-1.25) | 0.201 |
| IP-10 | 0.106 | 0.732 | 1.25 (0.95-1.64) | 1.04 (0.82-1.32) | 0.213 |
| SARS-CoV-2 IgG | 0.688 | 0.497 | 1.14 (0.61-2.11) | 1.23 (0.67-2.27) | 0.526 |
